# The LPP Indexes Baseline and Treatment-Related Changes in Anxiety Sensitivity

**DOI:** 10.1101/2024.10.23.24315611

**Authors:** Devin Butler, Asha Pavuluri, Spencer Fix, Norman B. Schmidt, Edward Bernat

**Affiliations:** University of Maryland College Park; Florida State University

## Abstract

Development of neurophysiological treatment targets has been an important way to validate and advance treatment implementations. Anxiety Sensitivity (AS), described as a fear of fear, is one of the most well-researched transdiagnostic risk factors for psychopathology and has been demonstrated to be associated with a number of different INT symptoms and disorders, making it an ideal intervention target for treatment of anxiety and mood pathology. Currently, there exists a gap in terms of understanding underlying neural mechanisms of AS and treatment-related change in AS. The current study sought to examine the impact of 2 brief computerized interventions on AS in order to assess the P300 and late positive potential (LPP) event-related potential (ERP) components as an index of both baseline and treatment change in the transdiagnostic anxiety sensitivity construct. Participants received one of the following, with a 1-month follow-up: an anxiety reduction intervention of cognitive anxiety sensitivity treatment (CAST) alone, a mood intervention of cognitive bias modification interpretation (CBM-I), a combined CAST and CBM-I intervention, or a repeated contact control group. Baseline AS was significantly related to baseline P3/LPP, such that higher P3/LPP amplitude (heightened emotional reactivity) is associated with higher AS at intake. Further, treatment-related reductions in AS were associated with higher baseline reactivity: greater AS reductions at month one were associated with higher P3/LPP activity. Heightened emotional response was predictive of better treatment outcomes. Analyses also showed evidence for different mechanisms associated with CAST and CBM-I groups. Specifically, the CAST group evidenced predictions of treatment outcome in parietal areas, where the CBM-I group evidenced predictions in frontal areas. The Combined group evidenced both frontal and parietal activations, consistent with combining the effects from the CAST and CBM-I groups. These findings suggest that the P3/LPP is an index of cognitive-affective processes underlying AS at baseline and has predictive ability in assessing AS treatment outcome in that our observed treatment types engage different neural mechanisms.

## Introduction

Anxiety psychopathology is incredibly pervasive in society: over 40 million Americans live with an anxiety disorder, and it is the most common mental health concern in the United States with a prevalence rate of 19%, while also being the most prevalent mental disorder in Europe at 14%. In Europe, it is estimated that anxiety disorders cost Europeans 74 billion Euros annually (Gustavsson et al., 2011; National Alliance on Mental Illness, 2024; H. U. Wittchen et al., 2011). With such high prevalence rates, the effect on both society and individuals are substantial. Anxiety Sensitivity (AS) is a transdiagnostic construct, which has been demonstrated to relate to a range of internalizing problems (anxiety, depression, and post-traumatic stress), as well as substance use). Research over the past four decades has identified AS as a causal risk factor, demonstrating that targeting AS in interventions can reduce symptoms across the range of related disorders. Recent work has synthesized work with AS interventions into a computerized and automated intervention, the Cognitive Anxiety Sensitivity Treatment (CAST). While there is now substantial evidence that CAST can reduce AS and related symptoms, there is a gap relative to understanding underlying neural mechanisms involved. These mechanisms could serve as treatment targets, or to assess change, and aid in treatment outcomes for such a widely prevalent issue such as anxiety. In the present project, we assess the P300 and late positive potential (LPP) event-related potential (ERP) components as an index of both baseline and treatment change in the transdiagnostic Anxiety Sensitivity construct after these brief technology-based interventions.

### Anxiety Disorders: Real-World Implications and Importance of Treatment

Anxiety is one of the most common and prevalent mental health problems in the United States as well as Europe (Kessler et al., 2012; Schiele et al., 2021; H. U. Wittchen et al., 2011). Financially, anxiety disorders regularly cost billions annually worldwide in terms of direct and indirect costs (Gustavsson et al., 2011). Anxiety pathology is a significant factor that, through strong association with debilitating symptoms, high medical costs, and suicidal ideation and behavior, necessitates a great deal of individual and societal burden (Stanley et al., 2018). Anxiety, and its associated symptomatology, affects work productivity, daily livelihood, and overall well-being globally. Thus, it is vital to develop a low-burden, neuroscience-informed transdiagnostic treatment in order to have a high impact generalizable to many. Such treatments may be especially impactful for those that identify with marginalized populations (e.g., racial/ethnic groups, low-socioeconomic status, substance use, and many other stigmatized identities) who are generally less likely to access and receive traditional treatment.

### Importance of Neurophysiological Treatment Predictors

Many individuals do not respond to currently available treatments for anxiety and mood disorders, with some response rates estimated at approximately 60% (Loerinc et al., 2015; Nimatoudis et al., 2004; Pollack, 2001a; Roth-Rawald et al., 2023). There is a need for more effective treatments as well as more informed and individualized treatment selection (Simon & Perlis, 2010). As remission rates for anxiety remain minimal even after treatment (H.-U. Wittchen, 2002), identifying objective pre-treatment measures that predict participant treatment response is critical.

Identifying pretreatment measures such as neurophysiological correlates to transdiagnostic constructs known to relate to anxiety and mood disorders, like Anxiety Sensitivity (AS), would provide a more robust understanding of determining factors in treatment success. A number of sociodemographic and clinical measures have been identified to predict treatment outcome: a history of failed treatments (Ellis et al., 2014), pre-treatment symptom severity (Connor et al., 2001; Doehrmann et al., 2013; Karatzias et al., 2007; Otto et al., 2000; Tükel et al., 2006), comorbidity with other disorders (Baer et al., 1992), and avoidant personality traits (Chambless et al., 1997). Objective neurophysiological markers of a construct such as AS would be complementary to this kind of research; neurophysiological correlates could offer a deeper insight into underlying mechanisms of internalizing disorders, and could influence future interventions.

### Anxiety Sensitivity: A Well-Researched Transdiagnostic Treatment Target

Anxiety Sensitivity (AS) is one of the earliest transdiagnostic constructs, first developed by Reiss (Reiss et al., 1986), indexing a “fear of fear.” More specifically, AS is defined as the extent to which a person is fearful of anxiety-related sensations due to intense beliefs that these sensations will have negative social, physical, and/or cognitive ramifications. Individuals high in AS have a higher tendency to misinterpret symptoms of anxiety as a sign of real and impending danger(Schiele et al., 2021), and through this misinterpretation they experience excessive worry and intensified anxiety, allowing AS to be seen as an “anxiety amplifier” that creates a cycle of anxious despair for an individual (Ghisi et al., 2016).

AS is one of the most well-researched, widely-accepted, and validated transdiagnostic constructs. Transdiagnostic models of psychopathology describe latent dimensions theorized to underlie and cause related disorders, and AS can be understood in the context of one of the primary current transdiagnostic models of psychopathology broadly, the hierarchical topology of psychopathology (HiTOP) model (Kotov et al., 2017). HiTOP represents psychopathology in 3 latent dimensions: internalizing (INT; depression, anxiety, and posttraumatic stress), externalizing (EXT; antisocial behavior, substance use, and dysregulated behavior), and thought disorder (TD; psychotic-thinking related disorders), with a newer overall measure of general psychopathology (GP; representing shared variance across INT, EXT, and TD).

Anxiety Sensitivity is a good intervention target for treatment of anxiety and mood pathology, as it is a causal risk factor for both (Naragon-Gainey, 2010). AS has been demonstrated to be associated with a number of different internalizing (INT) symptoms and disorders (e.g. fear, anxiety, depression, PTSD), as it prospectively predicts clinical diagnosis of related psychopathology such as anxiety, depression, panic, and posttraumatic stress symptoms (Schmidt et al., 1997, 2007; Sturges et al., 1998). AS not only predicts anxiety and mood disorder onset, it also plays a fundamental role in the maintenance of these INT problems (Ghisi et al., 2016). AS has been seen to be elevated across anxiety diagnoses, suggesting it is central to the phenomenology of anxiety disorders (Olatunji & Wolitzky-Taylor, 2009). AS is well-positioned as a transdiagnostic intervention target, as it is not only associated with diagnosis and maintenance, but also to treatment: treatment for AS has been demonstrated to reduce AS, and that reductions in AS produce reductions in current and prospective INT symptoms (Forsyth et al., 2000; Sturges et al., 1998).

With so much research on AS and how individuals high in AS process incoming stimuli, there is little established knowledge on cognitive processes underlying AS and the mechanisms underlying the relationship between AS and information processing. Of the few studies published on the topic, inconsistencies in methodology and findings limit conclusive evidence (Saulnier et al., 2021; Stevens et al., 2018). As AS has implications for a larger number of INT problems, research into objective measures of exploring underlying processes is crucial in ameliorating symptoms for millions worldwide.

### Cognitive Anxiety Sensitivity Treatment (CAST): A Suitable Treatment for AS

CAST is a brief and easily-implemented technology-based intervention that has proven to be effective in modifying AS: more specifically in targeting reductions in AS posttreatment (Schmidt et al. 2014). CAST is fully-automated, as it does not require therapist or experimenter participation, with two core components: psychoeducation and interoceptive exposure (IE) (Schmidt & Trakowski, 2004). The psychoeducation section provides corrective information regarding the nature of symptoms brought on by stress and anxiety (e.g., elevated heart rate, difficulty concentrating). Individuals are taught (using audio and video presentations, as well as interactive features) that physiological arousal and bodily sensations resulting from stress/anxiety are not dangerous, and that their elevated level of AS suggests they may have developed a conditioned fear of this arousal. This conditioned fear is then modified through the use of repeated IE exercises (i.e., computer guided hyperventilation) to attenuate the fear response to these bodily sensations. Several additional IE exercises are described and participants are encouraged to complete on their own.

CAST is a robust treatment for AS, with research showing AS is modifiable using CAST, such that posttreatment reductions in AS show reductions in anxiety and mood pathology (Capron et al., 2017; Schmidt et al., 2007, 2014, 2017). After just one session of CAST, nontreatment seeking participants with elevated AS can produce significant reductions in AS at posttreatment, relative to controls (Schmidt et al., 2014). CAST is an efficient treatment for reducing AS, and consistent with the transdiagnostic nature of AS and thus CAST, it is shown to have broad beneficial outcomes, as it has been shown to reduce a variety of INT symptoms, including anxiety, PTSD, depression, insomnia, and suicidal ideation(Mitchell et al., 2014; Raines et al., 2015; Short et al., 2015).

Culminating transdiagnostic approaches with constructs such as AS through a low-burden and effective technology-based delivery such as CAST supports the goal of developing more coherent interventions with strong impact. CAST has been shown to reduce AS, and to maximize the potential for effective treatments for those suffering from anxiety-related symptoms, utilizing CAST in the assessment of underlying neural mechanisms of AS is needed. In order to truly refine our understanding of cognitive processes underlying AS, an effective and efficient treatment designed to modify AS must be placed within that context. Measuring neurophysiological indices both pretreatment and posttreatment will provide evidence for correlates, and thus evidence for identifying future treatment targets.

Overall, due to the automated nature and core focus on the transdiagnostic AS construct, CAST has strong potential for use in low-resource environments, reducing AS and thus associated INT problems. Thus, there are strong opportunities to assess cognitive-affective mechanisms underlying change in AS, as was done in the present study, through the use of event-related potentials (ERPs).

### ERP Indices of Neurophysiological Processes

With the extensive behavioral work done developing and implementing CAST, including mechanisms of change, there are strong opportunities to extend this work to understand neurophysiological mechanisms that underlie these processes. Validation of the underlying neural mechanisms is innovative and can provide both knowledge relative to mechanisms and potential new neurophysiological treatment targets.

EEG event-related potential (ERP) methodology is well-suited to assess the constructs and relationships in this current study, with a number of advantages to the techniques including, but not limited to: they directly reflect neural activity, have excellent temporal precision to index early and rapid changes in neural processes, and specific components show particular responses to different types of stimuli, allowing for precise inferences of the core processes they index (Hajcak et al., 2010). Further, research has already supported EEG/ERP analyses as validated techniques of finding neural correlates to transdiagnostic constructs, including neural correlates of anxiety (Bernat et al., 2020; Hall et al., 2007; McTeague et al., 2020; Sha et al., 2019; Sharp et al., 2015). In work focused on modulating anxiety and mood symptoms, validated and clinically translatable measures of emotional processing, reactivity, and regulation are vital to producing novel research and replicable findings. There is a demonstrated record of using ERP measures from task protocols to index transdiagnostic measures. This work is evidence of the effectiveness of utilizing EEG and ERP measures in order to adequately assess the transdiagnostic concepts employed in this study.

Importantly, the two measures of interest in this study, the ERP components P300 and Late Positive Potential (LPP), have been used extensively in assessing the processing of emotional stimuli (Hajcak et al., 2010). The P300 is defined as a broad positive deflection maximal between 300-500 milliseconds post-stimulus onset (Hajcak et al., 2010; Johnston et al., 1986; Polich, 2007). The LPP follows the P300 and is much more sustained, with some characterizations of average activity defined from multiple timepoints until 2000ms post-stimulus presentation(Cuthbert et al., 2000; Foti & Hajcak, 2008). They are both commonly assessed from picture viewing and emotion regulation tasks (Bernat et al., 2011; Hajcak et al., 2010). Both the P300 and LPP show increased positivity following emotional stimuli relative to neutral stimuli, and work further supports the prevailing notion that affective modulation of the LPP measure is quite typical (Bernat et al., 2011; Cuthbert et al., 2000; Schupp et al., 2000). Increased positivity to emotional stimuli during the P300/LPP is described as a marker of motivated attention/incentive salience, and Bradley and colleagues defined motivated attention as the notion that emotion directs attention and guides subsequent processing (Bradley et al., 2003). The P300/LPP showing a larger positive peak to emotional stimuli relative to neutral reflects, in part, automatic processing of emotional stimuli due to intrinsic motivational significance.

This study assesses the P300 and LPP as indices of both baseline and treatment change in the transdiagnostic AS construct. The study integrates transdiagnostic approaches, including the intervention target (AS), psychopathology measures (INT problems), and underlying neural mechanisms in order to assess the utility, and underlying mechanisms, of an established transdiagnostic AS treatment (CAST) in reducing AS and INT symptoms. This study sought to relate AS to underlying neural mechanisms, as given what is known about AS, it may better reflect underlying systems and serve as evidence for empirical and objective referents in cognitive-affective processing. Affective responding in a picture-viewing and emotion regulation task was assessed, as disruptions in emotion regulation have been associated with various forms of psychopathologies, including aforementioned INT problems such as anxiety disorders, thus centering these disruptions as a central part of psychopathology (Aldao et al., 2010; Gross & Jazaieri, 2014; Gross & Muñoz, 1995; Sheppes et al., 2015). While there are many approaches to assessing affective responding, this study utilized a variant of common picture viewing/regulation task which has been well-validated using ERP measures (Bernat et al., 2011; Krompinger et al., 2008; Moser et al., 2006). Overall, the study will relate AS to underlying neural mechanisms, as given what is known about AS, it may better reflect underlying systems and serve as evidence for empirical and objective referents in cognitive-affective processing.

### Addressing Gaps in Literature on AS and Neurophysiological Measures

Altogether, this study addresses a need in the literature for more research as it relates to neurophysiological mechanisms and correlates of anxiety treatment outcomes. Development of neurophysiological treatment targets has been an important way to validate and advance treatment implementations. Currently, there exists a gap in terms of understanding underlying neural mechanisms of AS and treatment-related change in AS. As pointed out by Salunier and colleagues (Saulnier et al., 2021), cognitive processes underlying AS are poorly understood, and these processes can be nicely elucidated by ERPs. Though AS is well-researched and understood as a transdiagnostic risk factor for multiple forms of psychopathology (Naragon-Gainey, 2010), there exists little research on its underlying mechanisms, and using event-related potentials such as the P3 and LPP provide useful tools for refining our understanding of any core processes. The current study serves to mitigate the dearth of knowledge in the literature on such a well-studied transdiagnostic construct in hopes to broaden its impact as a treatment target.

Research has suggested that transdiagnostic psychopathology constructs have direct referents in neuroscience (McTeague et al., 2020; Sha et al., 2019), but these meta-analyses focused on whole-brain neuroimaging or resting-state fMRI. The current study evaluated treatment on a construct that is already proven to be productive and fruitful in our understanding of anxiety pathology. Validating underlying neural mechanisms is innovative in that it provides a possibility of neuroscience-informed intervention for anxiety and mood pathology. Anxiety disorders and symptomatology is widespread issue for millions worldwide. This study leveraged a validated transdiagnostic construct (AS) with a treatment proven to be effective in reducing it (CAST) and utilized ERPs to identify neurophysiological indices that can advance future interventions and benefit more individuals overall.

### Current Study

The primary aim of this study is to examine the utility, and underlying neural mechanisms, of an established transdiagnostic AS treatment (CAST) in reducing AS. Through measuring AS at baseline and at a 1-month post-intervention follow up, the current study can examine underlying cognitive affective processes in individuals elevated in AS, as well as the efficacy of treatments targeting AS, with aims to see AS reductions. We hypothesize P3 and LPP measures are associated with pretreatment baseline AS scores, as well as a P3/LPP sensitivity to change scores between treatment groups. Such a finding would advance our understanding of neural mechanisms for cognitive-affective processing, as it would show AS is capable of being measured physiologically, and that an ERP measure is a vital tool for understanding these mechanisms and processes.

## Methods

### Participants

Participants (N = 263) were recruited from the community to participate in a computerized treatment study targeting risk factors relevant to Anxiety Sensitivity (AS). Participants’ ages ranged from 18 to 79 years old, average age 35.89 years, 148 females. All participants were 18 years of age or older and were screened for neurological conditions, visual impairments, and/or traumatic brain injuries.

Other inclusionary criteria were partially based on elevations on AS. The threshold was set to above average risk on Anxiety Sensitivity’s cognitive concerns subfacet (>8) (Schmidt & Joiner, 2002). Other inclusionary criteria included: English-speaking participants; no evidence of uncontrolled psychotic-spectrum or bipolar disorders; no imminent suicide risk; no significant medical illness (e.g., cardiovascular disease) that would prevent the completion of interoceptive exposure exercises (i.e., repeated induction of bodily sensations); and/or not participating in psychotherapy at intake. Participants were allowed to be on medications if they were on a stable dose for at least six weeks. Education levels ranged from less than high school, high school, some college, four-year college degree, and graduate school or higher. The racial breakdown ranged as follows: Caucasian, African American, Asian, Native American, Pacific Islander, and other (e.g., biracial). Additionally, some of the sample identified as Hispanic.

### Missing Data

Participants were recruited approximately evenly into the ANX/CAST (n = 65), MOOD/CBM-I (n = 66), Combined (68), and Control (n = 64) groups at baseline. All participants were included in analyses across conditions at baseline, as participants were randomly assigned to each condition and there had yet to be any treatment conducted.

Across conditions, 44 people missed their 1-month post-intervention session: 10 participants in the CAST group, 13 in the CBM-I group, 10 in the Combined group, and 11 in the Control group. Thus, measurements of AS were not conducted for these participants, and total participants for analyses at 1-month post treatment amounted to 219. Subsequent analyses that include M1-BL treatment change scores excluded these participants. Further, investigation into treatment effects focused on treatment groups, relative to the control group, leaving the sample size for this set of analyses to be 166 across the 3 treatment conditions.

### Procedures

#### Screening Appointment

All study procedures were approved by the university’s institutional review board. After meeting initial inclusionary criteria through a brief telephone interview, individuals were brought in to complete a screening appointment. The screening appointment was meant to be more intensive. Suicide risk was assessed at all timepoints (via interview and suicide-relevant self-report measures), and based on risk designated for an individual, actions were in place to provide appropriate, standard care (e.g., suicide hotline information, creating a safety plan, means restriction; Chu et al., 2015). During this screening appointment, along with the thorough suicide risk assessment, participants also underwent a diagnostic interview (First et al., 2016) with a trained therapist (Joiner Jr. et al., 1999). Participants then completed self-report measures to help determine study eligibility and inform diagnostic decisions. If, for any reason, participants were deemed ineligible based on the screening appointment, they were thanked for their time and given relevant community referrals based on their needs.

#### Baseline appointment

Participants completed a baseline neurophysiology assessment (EEG) and were provided with commensurate monetary compensation. *Intervention Appointments (3 sessions).* Participants received the intervention at a rate of one 60-minute session per week for three weeks, with all sessions being completed in the clinic. During each session, participants completed their assigned intervention, as well as subsequent assessment measures. In the active treatment conditions, Session 1 included the relevant psychoeducation and CBM interventions. Sessions 2 and 3 included only CBM. *Follow-up appointment (Month 1)*. Participants completed study questionnaires in an individual testing room. Upon completion of the measures, participants were scheduled for their next follow-up appointment (1 month after baseline/post-treatment) and were awarded any monetary compensation they earned. *Randomization*. Eligible participants were randomized, using an online random number generator, to one of four possible study conditions.

### Intervention Procedures

#### Overview

The three treatment groups received a computer-based intervention that combined psychoeducation, brief exposure therapy (CAST), and Cognitive Bias Modification-Interpretation (CBM-I). The psychoeducation component lasted approximately 45 minutes in the first session and focused on the nature of stress and the effect it has on the body. Participants were taught that the physiological arousal associated with stress is not harmful to them, and they were instructed to participate in guided exercises to correct the fear response associated with said arousal. CAST functioned as the guided exercise. CAST consists of a psychoeducation and exposure components. During the CAST portion of the session, participants were first directed to complete a standardized assessment of their fear to different arousal sensations. Participants completed repeated exposure trials engaging in an arousing sensation, such as hyperventilation, and subsequently rated the level of arousal they experienced during the exercise (on a scale of 1-10). They were told they would repeat each exercise until their subjective rating of distress was rated as minimal (i.e. a rating of 0-1). They were also instructed to complete one set of each of the exercises daily until none of the exercises generated any fear/distress.

CBM-I focuses on changing an individual’s automatic or reflexive interpretation of incoming information by providing feedback to participants about whether this interpretation of stimuli was correct. During the task, participants were presented with a word (e.g., “excited”) for 1 second, followed by the presentation of a sentence (e.g., “You notice your heart is beating faster”). They were subsequently asked to, by button press, determine whether they thought the word was related to the sentence or not by pressing “yes” or “no.” On half of the trials, the word and sentence combination created a benign meaning (aforementioned example), while the other half of trials created an anxious meaning (e.g., “stressful” followed by “Your mind is full of thoughts”). Participants were given feedback during training: “correct” feedback was defined as judging the anxious combinations to be “unrelated” and the benign combinations to be “related”, while feedback was defined as “incorrect” if they judged the anxious combinations to be related and the benign combinations to be unrelated, they were given feedback that the response is “incorrect.” For incorrect responses, the feedback was also accompanied by a horn blast (approximately 85 decibels). An interpretation bias is thus typically measured by the number of trials in which participants endorse benign relationships and reject anxious or depressed combinations. Participants completed 40 test trials with no reinforcement (incorrect or correct feedback), followed by 80 training trials in which each response was given feedback. After the test trials, participants then took a short 5-minute break, and during this break they completed a filler task (simple math problems), followed by another 80 training trials. Finally, they were given 40 more test trials of novel words and sentences that they had not seen before.

### Treatment Groups

#### Anxiety Intervention Condition (ANX)/ CAST Group

Participants who received the anxiety intervention completed CAST (Schmidt et al., 2014) and an AS-focused CBM program (CBM-I for AS) (Capron & Schmidt, 2016). As mentioned previously, CAST is a fully computerized, 45-minute intervention designed to model the techniques, both educational and behavioral, that are commonly used in anxiety treatments. The CBM-I component used in this intervention condition was programmed using E-Prime software (Schneider et al., 2002). The trial sequence for the CBM-I component is described in detail above in the **Overview** section.

#### Mood Intervention Condition (MOOD)/ CBM-I Group

The mood condition paralleled the anxiety condition in that it included a top-down psychoeducational portion as well as a bottom-up CBM portion. Participants who received the mood intervention completed a fully computerized, 50-minute intervention designed to model the educational and behavioral techniques that are commonly used in the treatment of mood disorders, which is the same tact delivered in the ANX condition, except there the techniques being modeled are those that treat anxiety disorders. In the mood intervention, participants were taught that negative beliefs about being isolated and burdensome are usually inaccurate. Following this mood intervention, behavioral activation techniques were introduced as a way to decrease isolation and feelings of burdensomeness. The CBM-I component of the Mood Intervention Condition was developed by Holmes and colleagues (Holmes et al., 2006). 100 scenarios were presented across five training blocks, containing 20 scenarios each. For more information regarding the CBM-I used, see Holmes et al. (2006).

#### Combined Intervention Condition (COMBINED)

Participants assigned to the combined condition completed both the anxiety and mood intervention conditions. Thus, while this meant that the Combined group intervention was not matched for length, it was still delivered over three sessions, consistent with the other treatment conditions. The order of the mood and anxiety interventions, as well as their respective CBM tasks, were counterbalanced across participants at each session.

#### Repeated Contact Control Condition (RCC)/ Control Group

A relatively simple repeated contact intervention has been suggested to be effective in reducing suicide (Fleischmann et al., 2008; Motto & Bostrom, 2001). Therefore, a repeated contact control condition, or RCC, represents an ethically justifiable control in studies like this proposed one, in which participants at risk for suicide and AS are used. At their baseline appointment, participants in this RCC condition were assigned a personal study coordinator. Participants met with their study coordinator once per week for three weeks (corresponding to the treatment session intervals for those in the active treatment conditions) for a brief check-in where suicide risk was evaluated and preventative measures were taken if needed (e.g., safety plan, lethal means counseling, resource recommendations).

### Diagnostic interview

#### Structured Clinical Interview for DSM-5, Research Version (SCID-5-RV)

The SCID-5-RV is a semi-structured clinical interview that assesses the presence of DSM-5 psychiatric diagnoses (First et al., 2016). The SCID-5-RV was administered by clinical psychology doctoral student therapists who underwent a systematic training procedure. Therapists only began conducting diagnostic interviews once they demonstrated high levels of reliability. All diagnostic decisions were reviewed by a licensed clinical psychologist to ensure high levels of diagnostic accuracy. Diagnostic raters were blind to experimental conditions.

### Self-Report Measures

#### Anxiety Sensitivity Index-3

The primary treatment outcome measure was the Anxiety Sensitivity Index (ASI-3) (Taylor et al., 2007), which is an 18-item self-report measure designed to assess an individual’s tendency to interpret anxiety-related sensations as potentially harmful or dangerous. These sensations may take the form of thoughts/cognitions, physiological experiences, or social situations. The ASI-3 has strong psychometric properties. In the present study, the ASI-3 cognitive concerns subscale demonstrated excellent internal consistency (α = .94) at baseline with scores ranging from 0-24 at baseline. Example items include: “When I have trouble thinking clearly, I worry that there is something wrong with me” (cognitive), “It scares me when my heart beats rapidly” (physical), and “It is important for me not to appear nervous” (social). Respondents use a 5-point Likert-type scale ranging from 0 (very little) to 4 (very much) to indicate the extent to which each item reflects their typical experience. In the present study, ASI was tracked at baseline appointment (BL), as well as one month post-treatment (M1)

### Experimental Procedures

Prior to treatment condition randomization, participants completed a structured clinical interview and self-report measures prior to being scheduled for the EEG session. The recording session consisted of several different tasks, including the emotional picture paradigm (presented second after a resting task), which was utilized for the current study. All data presented in the current study were collected prior to treatment randomization. After participants completed the baseline questionnaires and psychological assessments, they participated in baseline EEG measurements, as well. These measurements included the following tasks: Resting, Emotion Regulation, Fast Pictures, Go/No-go, Gambling, and Oddball.

#### Psychophysiological Data Collection

All neurophysiological data was collected in a dimly lit sound attenuated room, where E-prime version 2.0 was used to present the computer tasks. Experimental stimuli was presented on a 21-inch Dell high-definition CRT color monitor, centrally placed in front of participants at a viewing distance of 100 cm.

Neurophysiological data was recorded using a BrainVision 96-channel actiCap (sintered Ag-Ag/Cl; international 10-20 system;(Jasper, H.H., 1958)) as well as a 24-bit battery-supplied active channel amplifier. Horizontal electrooculogram activity was recorded from electrodes placed on the outer canthus of both eyes, while vertical electrooculogram activity was recorded from electrodes placed above and below the left eye. Impedances were kept below 10 kΩ. EEG signals were vertex referenced (FCz) during recording. Recordings were collected using a 500Hz sampling rate, analog 0.05 to 100Hz bandpass filter, and digitized at 1000 Hz using BrainVision PyCorder (Brain Vision LLC).

### Data Processing

#### Preprocessing

The EEG data was preprocessed using custom Matlab scripts based on EEGLAB (Delorme & Makeig, 2004), and analyzed using the Matlab-based Psychophysiology Toolbox (PTB) (Buzzell et al., 2022). The preprocessing steps are set up to clean and correct the EEG datafiles. In the end, there will be a set of preprocessed mat files that can then be analyzed through the PTB. Pre-processing included downsampling to 512 Hz, bad channel detection and replacement, epoching, and eye-blink removal (Gratton, 1998). Bad channels were identified as having activity four standard deviations away from the mean of all other non-ocular channels. These channels were replaced using the mean of surrounding electrodes. This method, used previously (Anderson et al., 2015; Maurer et al., 2016; Steele et al., 2014, 2015, 2016) was implemented to identify very large artifacts and remove them from the data before applying more stringent data cleaning steps in post-processing. Epochs of 3,000ms were taken from 1,000ms pre to 2,000ms post-stimulus with a 500ms pre-stimulus baseline correction, 150ms post-stimulus baseline correction, and were re-referenced to averaged mastoid sites. The respective triggers for each task were set in the respective task preprocessing script (e.g., loss and gain trials in the gambling task).

### Emotion-Regulation and Picture-Viewing Paradigm

We closely replicated the instructions used first by Jackson et al. (Jackson et al., 2000). The aim was to encourage participants to use cognitive reappraisal strategies as articulated by Gross (1998). In short, participants were told that before a picture was presented, they would be instructed to enhance, suppress, or view the emotion they felt toward the picture. To enhance the emotion, they were asked to increase the intensity of emotion they felt. To suppress the emotion, they were instructed to decrease the intensity of emotion they felt. Suppress, as operationalized here (and in Jackson et al., 2000), corresponds to a reappraisal strategy (cf. Gross, 1998). In addition, it was explained that participants would sometimes be asked simply to view pictures, in which case they were not to attempt to manipulate their emotions. In all cases they were advised to stay focused on the picture and the induced emotion. Incorrect methods of regulation were described as generating unrelated emotions, thinking of things unrelated to the picture, looking away from the picture, or only focusing on parts of the picture.

The emotion regulation task is based on images from the International Affective Picture System (Lang et al., 1997). Participants were shown 24 pleasant pictures (i.e., socially rewarding or erotic pictures) (Kanske et al., 2013), 24 unpleasant pictures (i.e., threat/mutilation images), and 60 neutral images (i.e., non-affective people, places, or objects). Emotional images were divided into six blocks (AS, unpleasant, pleasant, thwarted belongingness, perceived burdensomeness, and suicide). Three instruction types were utilized for pleasant and unpleasant images: View (i.e., passively observe the image), increase (i.e., enhance emotional response), and decrease (i.e., reduce emotional response). On view trials, participants were instructed to allow their emotions to happen naturally, and to not manipulate their emotions in any way. For the increase and decrease trials, participants were instructed not to focus on any specific part of the image or avert their gaze from the picture in order to regulate their emotional reaction and to instead use other emotional regulation strategies. The order of the presentation of the instruction blocks (i.e., which instruction type was presented first, second, and third), images, blocks, and sub-blocks were counterbalanced across participants. For neutral images, only view trials were conducted, to avoid confusion in trying to regulate emotional responses to non-emotional images. In line with prior work (Moser et al., 2006), each trial began with a regulation trial-type cue (i.e. Increase, Decrease, or View) lasting 2000ms, which was followed by the image presentation for 6000ms. Inter-trial intervals consisted of a blue fixation point presented for 2000ms. Analyses of reactivity in this study only included unpleasant (UP) imagery, in line with previous work (Allan et al., 2019; Jackson et al., 2000).

### Data Reduction

The time-domain P3 ERP component was defined and used for analysis of early processing differences between groups. The P3 was defined as the positive deflection ranging from about 281ms to 422ms (36 to 54 bins), as it is generally maximal around 350ms(Hajcak et al., 2010). Also assessed was the late positive potential (LPP), a late slow wave ERP component ranging from 350ms to 1500ms. Discussion of the LPP coarsely refers to the positivity beginning in and extending beyond the P300 ERP (Hajcak et al., 2010; Hajcak & Foti, 2020). Due to the similarities and overlap, it can be difficult to parse the P300 and LPP, thus the strong amount of overlap justifies looking at one component window, as we do in this study: the time window containing the P3 and LPP will be referred to as the P3/LPP. In their review, Hajcak and colleagues suggested to study and evaluate the P3/LPP at parietal sites and frontal/central sites. Thus, based on the scalp topography of the LPP in our data and the selection of centro-parietal electrodes in previous studies, we defined analyses across two clusters: a frontal cluster, and a parietal cluster (referenced together as FP). The frontal cluster was a subset of 14 electrodes, centered around a frontocentral electrode (FCz). The parietal cluster was a subset of 13 electrodes, centered around a parietal electrode (Pz) (Brown et al., 2012; Gable et al., 2015; Thiruchselvam et al., 2011).

The ERP measures will be utilized in this study because the transdiagnostic constructs addressed in this study have direct referents to neuroscience(McTeague et al., 2020; Sha et al., 2019). Merging our understanding of psychopathology and treatment with ERP measures could lead to a characterization of separable transdiagnostic systems underlying problem behavior.

### Data Analytic Plan

To assess hypotheses, means will be compared through t-tests and ANOVAs. A t-test will be conducted to compare AS at baseline to AS at 1-month post-treatment. We’re predicting here that AS will decrease, as seen in lower AS scores posttreatment, as further replication and validation of the treatment efficacy in targeting AS. A one-way ANOVA will be conducted to assess a relationship between baseline AS and baseline LPP measures. Baseline P3/ LPP measures will be correlated with measures of change in AS between baseline and 1-month (change score) in order to assess for CAST-related treatment change between groups. This will be done using GLM with a continuous predictor (AS change score) and a categorical predictor (group: Anx/Cast, Mood/CBM-I, Combined), with the P3/LPP measures as the dependent variable. A one-way ANOVA will be conducted to correlate ER task instruction to AS change score. This analysis will assess for any potential differences in treatment effects between groups across emotion regulation instructions.

## Results

### Baseline ERP Measures Significantly Associated with Baseline AS

To assess for prediction at intake, BL AS was correlated with BL P3/LPP measures. An ANOVA was conducted to assess a relationship between the P3/LPP ERP and baseline AS. Pearson correlation coefficients were computed across all participants, and across task instruction, at baseline to assess the linear relationship between ERP and AS. All significant correlations reported are significant at the *p* < .05 level.

ANOVA results revealed a significant effect of baseline AS on P3/LPP [F(1, 261) = 9.12, p = 0.003, ηp2 = .034] (**Table 1**). Across all experimental conditions, there does exist a relationship between baseline AS and the P3/LPP. Baseline AS was not related to topographical differences (frontal and parietal, or FP) [FP x AS: F(1,261) = .000, p = .989, ηp2 = .000], and AS was not related to P3/LPP differences [Component x AS: F(1,261) = .563, p = .454, ηp2 = .002]. ANOVA results indicate that a relationship between Anxiety Sensitivity and P3/LPP is present: higher P3/LPP amplitude is associated with higher BL AS.

**Table 1.**
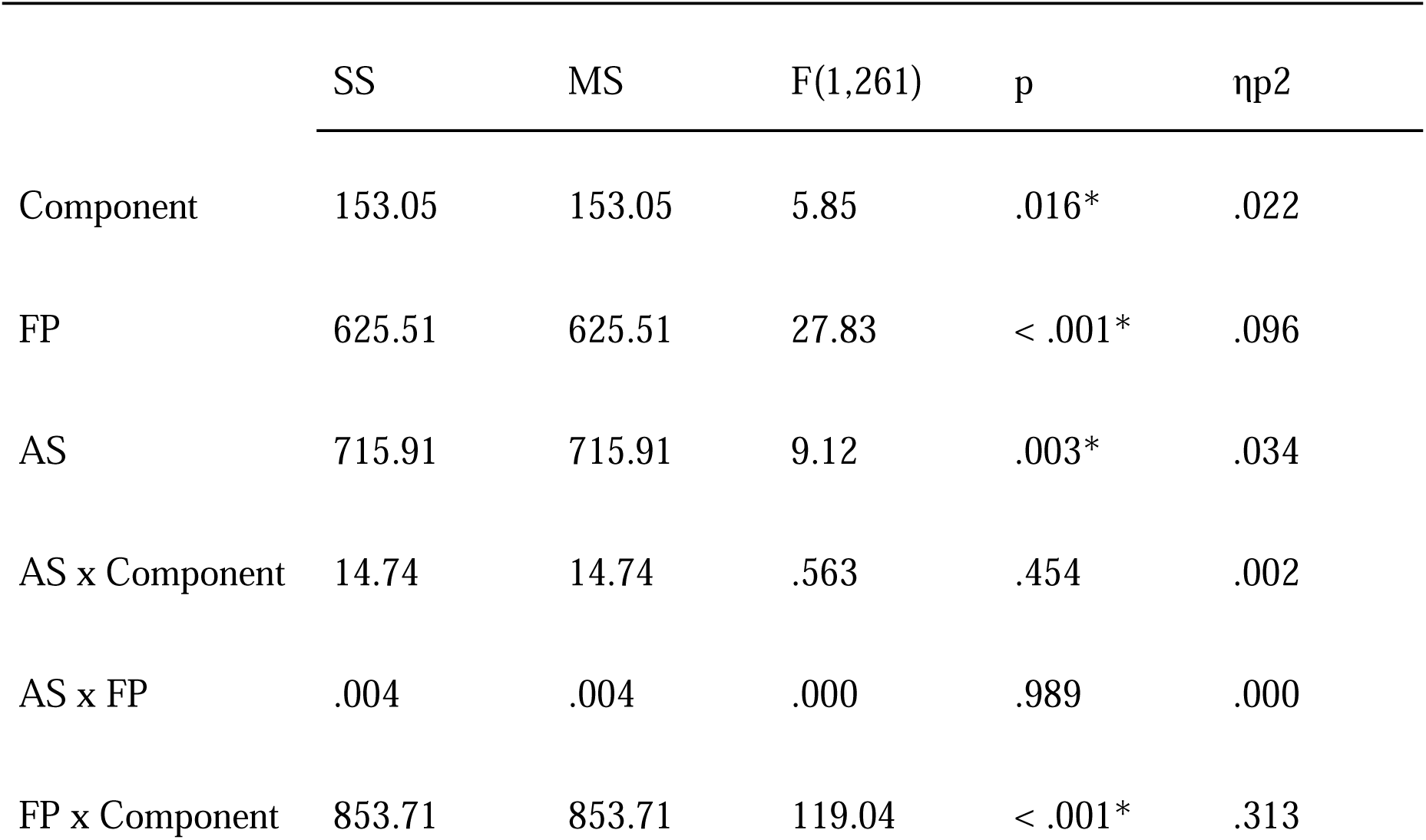
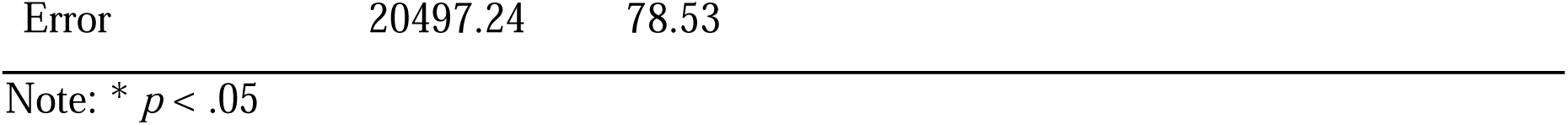
Omnibus ANOVA: Baseline AS and P3/LPP.

| | SS | MS | F(1,261) | p | $\eta^2$ |
| --- | --- | --- | --- | --- | --- |
| Component | 153.05 | 153.05 | 5.85 | .016* | .022 |
| FP | 625.51 | 625.51 | 27.83 | < .001* | .096 |
| AS | 715.91 | 715.91 | 9.12 | .003* | .034 |
| AS x Component | 14.74 | 14.74 | .563 | .454 | .002 |
| AS x FP | .004 | .004 | .000 | .989 | .000 |
| FP x Component | 853.71 | 853.71 | 119.04 | < .001* | .313 |

|  |  |  |
| --- | --- | --- |
| Error | 20497.24 | 78.53 |

Note: \* $p < .05$

Correlation analyses confirmed the nature of the relationship between BL AS and P3/LPP activity, such that higher LPP amplitude (heightened reactivity) is associated with higher AS at intake, *r(*261) = .184, *p* = .003. Taken together, these results support the hypothesis that P3 or LPP amplitude can be used to index a sensitivity to AS, such that higher levels of AS were associated with higher LPP amplitude.

### Relationship Between Change Scores and Task Instruction on P3/LPP Amplitude

To assess a sensitivity to change scores, this time taking into account task instruction, an ANOVA was conducted to correlate instruction to change scores. The omnibus ANOVA revealed that, across treatment groups and other experimental conditions, task instruction was not related to M1-BL change scores in terms of P3/LPP amplitude [F(1,164) = .191, p = .662, ηp2 = .001] (**Table 2**). Though there was emotion regulation, as seen by marginally significant relationship between task instruction and amplitude, due to the task instruction not being significantly related to AS change scores, further analyses conducted were collapsed across task instruction.

**Table 2.** ANOVA: Relationship between Instruction and Change Scores on Amplitude.

| | SS | MS | F | p | $\eta^2$ |
| --- | --- | --- | --- | --- | --- |
| Instruction | 27.80 | 27.80 | 2.79 | .097 | .017 |
| Instr. x $\Delta$ AS | 1.91 | 1.91 | .191 | .662 | .001 |
| Error | 1634.12 | 9.96 |  |  |  |

### Treatment-related Reductions in AS

To assess for treatment-related reductions in AS, both within and across treatment groups, paired samples t-tests were conducted to compare AS at baseline and AS 1-month after treatment (see **Table 3** for results).

**Table 3.** T-tests for Baseline and M1 AS within group and across treatment groups.

| Group | Baseline |  | 1-Month |  | *t* | *p* | Cohen's *d* |
| --- | --- | --- | --- | --- | --- | --- | --- |
|  | *M* | *SD* | *M* | *SD* |  |  |  |
| CAST | 31.62 | 14.83 | 16.44 | 12.21 | 8.51 | < .001* | 1.15 |
| CBM-I | 25.58 | 12.74 | 17.77 | 12.86 | 5.50 | < .001* | 0.755 |
| Combined | 29.82 | 14.41 | 14.93 | 13.82 | 8.24 | < .001* | 1.09 |
| Across Tx Group | 29.06 | 14.08 | 16.35 | 12.97 | 12.66 | < .001* | .985 |
| Control | 30.02 | 14.28 | 22.72 | 15.10 | 4.14 | < .001* | .564 |

Note: \* $p < .05$

Across treatment groups, there was a significant decrease in AS 1-month post treatment (M = 16.35, SD = 12.97) compared to baseline AS (M = 29.06, SD = 14.08), t(164) = 12.66, p < .001, Cohen’s d = .985. In the CAST group, there was a significant decrease in AS in the month after treatment (M = 16.44, SD = 12.21) compared to AS measured at baseline (M = 31.62, SD = 14.83) , t(54) = 8.51, p < .001, Cohen’s d = 1.15. In the CBM-I group, there was a significant decrease from baseline (M = 25.58, SD = 12.74) to M1 (M = 17.77, SD = 12.86), t(52) = 5.50, p < .001, Cohen’s d = .755. Similarly, there was a decrease from baseline (M = 29.82, SD = 14.11) to M1 (M = 14.93, SD = 13.84) in the Combined group, t(56) = 8.24, p < .001, Cohen’s d = 1.092.

Interestingly, there was also a significant difference in the Control group, in that there was a significant decrease in AS from baseline (M = 30.02 , SD = 14.28) to M1 (M = 22.72, SD = 15.10), t(53) = 4.14, p < .001, Cohen’s d = .564. To assess differences in treatment effects, a one-way between subjects ANOVA was conducted to compare the treatment effects in the 4 treatment groups. There was a significant effect of Condition on AS M1-BL change scores at the p < .05 level for the 4 treatment groups [F(3, 215) = 6.52, p < .001, ηp2 = .083]. Post hoc comparisons using the Tukey HSD test indicated that relative to the Control group, treatment effects were larger in the CAST group. The mean treatment change score (M1-BL) for the CAST group (M = −15.2, SD = 1.69) was significantly larger than the Control group (M = −7.38, SD = 1.73). The Combined group’s treatment effects were also larger relative to the Control group, as the Combined group’s mean change score (M = −15.00, SD = 13.30) was larger than the Control’s. The CBM-I group (M = −7.81, SD = 1.73) did not significantly differ from the Control group in terms of treatment effects. Further, no significant differences were found between the CAST group and the Combined group. The effects in the CAST group were however larger than the effects of the CBM-I group, as indicated by higher treatment change in CAST. Further, the Combined group (M = −15.00, SD = 1.65) also showed significant differences with the CBM-I group. These results indicate that the CAST and the Combined treatment groups showed the highest effect on treatment-related change in AS scores from M1 to BL. More specifically, CAST and Combined groups led to greater reductions in AS relative to the CBM-I and Control groups, while the CBM-I and Control groups did not differ in their treatment related AS reductions. See **Table 4** for a visual depiction of AS change from baseline to M1.

**Table 4.**
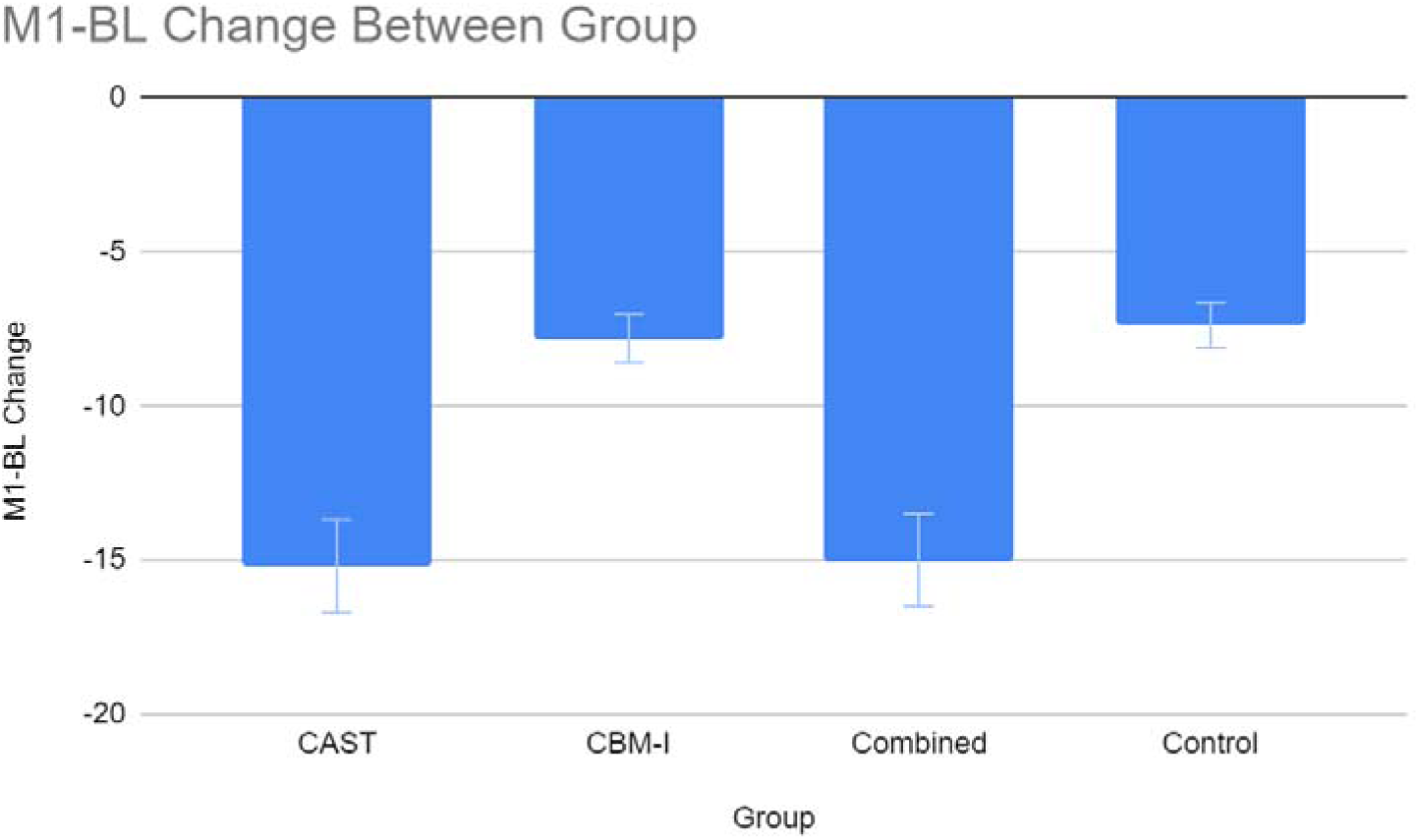
Change in AS from BL to M1 within Each Treatment Group and Control.

### AS Treatment Change predicted by BL P3/LPP Measures

Analyses were conducted to correlate baseline P3 and LPP measures with measures of change in AS between baseline and 1-month (change score). Results from ANOVAs with AS Change Score and treatment group as predictors (see **Table 5**), as well as correlation coefficients, are explained below. Results revealed a correlation between baseline AS scores, and treatment change scores from baseline to month one, r(166) = −.54, p < .01, indicating the higher AS scores were at baseline, the larger the reduction in AS at one month posttreatment.

**Table 5.** Omnibus ANOVA: M1-BL Change Score and P3/LPP.

| | SS | MS | F(1,164) | p | $\eta^2$ |
| --- | --- | --- | --- | --- | --- |
| Time | 503.36 | 503.36 | 17.47 | < .001 | .022 |
| FP | 1018.03 | 1018.03 | 43.79 | < .001 | .211 |
| $\Delta$ AS | 718.20 | 718.20 | 9.23 | .003* | .053 |
| $\Delta$ AS x Time | .488 | .488 | .017 | .897 | .000 |
| $\Delta$ AS x FP | .436 | .436 | .019 | .891 | .000 |
| FP x Time | 1417.91 | 1417.91 | 184.78 | < .001* | .530 |
| Error | 12763.65 | 77.83 |  |  |  |

Note: \* $p < .05$

Similar to results for the relationship between P3/LPP and baseline AS, omnibus testing revealed a significant relationship between AS change scores 1 month post treatment and baseline P3/LPP measures [F(1,164) = 9.23, p = .003, ηp2 = .053] (**Table 5**). This result indicates that across topography, and across the time windows, a relationship between baseline P3/LPP and change scores from baseline to one month is present. Across treatment groups, treatment-related effects are associated with baseline P3/LPP ERP measures. ASI change scores are not related to time [F(1,164) = .017, p = .897, ηp2 = .000], nor are they related to frontal and parietal topography differences [F(1,164) = .019, p = .891, ηp2 = .000]. Further, there exists an interaction between topography and time [FP x Time: F(1,261) = 184.78, p < .001, ηp2 = .530], stated earlier that parietal effects start earlier in time, at the P3, and frontal effects start later, during the LPP.

### Relationship Between AS Change Score and Treatment Group on P3/LPP Amplitude

To assess differential effects of treatment group on change score, an ANOVA was conducted (**Table 6**). The omnibus revealed that there were no significant differences between the 3 treatment groups on P3/LPP activity [F(1,164) = .670, p = .513, ηp2 = .008]. Regression analysis revealed that the relationship between P3/LPP activity and ASI change scores was not significant between treatment groups (Beta = .256, t = 1.11, *p* = .269). Due to effects not being significant between treatment group, further analyses collapsed across condition.

**Table 6.**
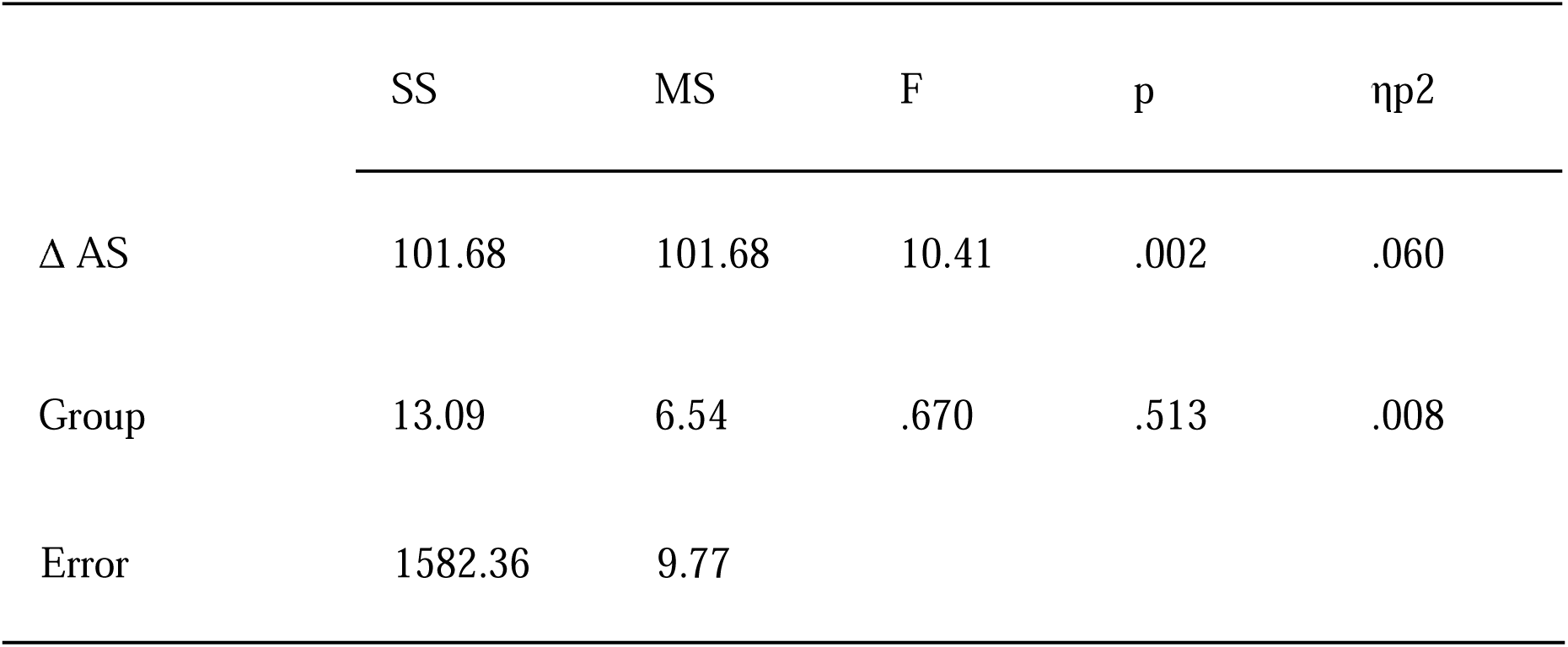
ANOVA: Relationship between Treatment Groups and Change Scores on Amplitude.

Correlation analyses reveal the nature of the relationship between treatment change scores and baseline P3/LPP activity. Importantly, baseline AS was correlated to AS treatment change across treatment groups, r(164) = −490, suggesting the higher the AS score at intake/greater the AS severity, the greater the treatment outcome. Across treatment groups, time windows, and clusters, ASI treatment change scores are correlated with treatment change, such that higher LPP baseline reactivity is associated with greater AS reductions across treatment groups, r(164) = −.247, p = .001. The control group did not show significant correlations between treatment change and EEG activity, r(51) = −.027, p = .846. Although the omnibus revealed no significant differences between treatment groups, a priori comparisons for AS Change Scores within Treatment Group in relation to P3/LPP amplitude were made below. These group assessments are partially justified by these correlation analyses, and motivated by the goal of assessing differences in the treatment groups, if more tentatively statistically due to the smaller N in each group.

#### Correlations Between M1-BL Change Scores Within Treatment Group and P3/LPP

Correlation analyses revealed significant associations within treatment group between AS change scores and P3/LPP amplitude, all within respective frontal and parietal clusters:

Within the CAST group, there existed a correlation between parietal P3/LPP amplitude and AS change scores, r(53) = −.269, p = .047, indicating that higher parietal P3 is associated with larger AS change scores, or greater AS reduction. Frontally, within the CAST group, there is no association between P3/LPP amplitude and change scores, r(53) = .012, p = .931.

To contrast these CAST group effects, correlation analyses revealed that across the entire P3/LPP window, within the CBM-I group, a similar relationship exists, but significant effects are found frontally: M1-BL AS change scores are associated with P3/LPP amplitude frontally: r(51) = −.293, p = .034, such that higher frontal P3/LPP amplitude is associated with greater AS reduction. Parietally, there is no such association within the CBM-I group across the time window, r(51) = −.201, p = .149.

Correlation analyses within the Combined group showed combinatorial effects. In both frontal and parietal clusters respectively, there existed an association between P3/LPP amplitude and treatment change scores. In the frontal cluster, amplitude within the Combined group was significantly correlated with AS change scores, r(56) = −.344, p = .008. There was a similar effect parietally, at marginal significance, r(56) = −.243, p = .066). These analyses showed that higher LPP amplitude was correlated with greater AS reductions, i.e. larger treatment change scores.

The control group showed no significant correlations between AS change score and P3/LPP amplitude (frontal: r(51) = −.057, p = .687; parietal: r(51) = −.065, p = .644).

Significance tests were then computed to assess for significant differences between groups in correlations between P3/LPP amplitude and AS change scores. In a Fisher Z test of significance between correlations coefficients representing the relationship between AS change score and both frontal and parietal P3/LPP amplitude in the CAST and Control groups respectively, analyses revealed a nonsignificant difference, (frontal: z = 1.098, p = .136. parietal: z = .324, p = .373). Similarly, for the CBM-I vs Control groups for frontal P3/LPP amplitude and AS change score, the difference was trend level, z = −1.296, p = .097, and the correlations were not significant parietally (z = −.309, p = .379). For comparisons between the Combined and Control groups, the correlations were significantly different in both frontal and parietal LPP activity (frontal: z = −2.154, p = .016. parietal: z = −1.558, p = .06). These tests of significant correlational differences suggest that the association between amplitude and AS reduction is uniquely parietal within the CAST group, frontal within the CBM-I group, and a combination of frontal and parietal within the Combined group when compared to Control.

As findings showed that the P3/LPP here was able to predict ASI treatment outcome (as observed by change score from baseline to month one), two regressions were run to assess if the P3/LPP predicted change in an Internalizing factor overall, as well as the extent ASI treatment explained this relationship. We assessed this in the combined group, where results demonstrated the strongest observed effect. First, a regression model with the ERP measure alone explained a significant amount of variance in the change in Internalizing from baseline to month one [F(1, 50) = 5.27, p = .026, R-squared = .095, Adjusted R-squared = .077], with P3/LPP activity as a significant predictor (Beta −.309, t = −2.39, p = .026). Negative direction in the P3/LPP estimate indicates higher amplitude is associated with greater treatment outcome/larger decreases in Internalizing from baseline to month one. To assess change in ASI as a candidate explanatory factor for the change in Internalizing, we added change in ASI to this regression as a predictor. Thus, this included both P3/LPP and the ASI change score from baseline to month one as predictors of change in Internalizing, the model is significant [F(1, 50) = 19.44, p < .001, R-squared = .257, Adjusted R-squared = .326], but this relationship becomes insignificant, such that P3/LPP no longer predicts INT change (Beta −.129, t = −1.06, p = .294), whereas ASI change score does (Beta .508, t = −4.41, p < .001), such that greater AS reduction is associated with greater internalizing reduction suggesting the relationship with P3/LPP is accounted for by AS, the target construct of our study.

## Discussion

### LPP Related to Baseline AS

The first main finding of this paper is a relationship between Anxiety Sensitivity and the LPP, such that higher amplitude, indexing heightened emotional reactivity, is associated with higher AS at intake. The LPP elicited from unpleasant images was related to AS. This relationship between AS and LPP is not entirely surprising, as previous literature has suggested AS is related to cognitive processes involved with emotional processing of stimuli, higher LPP to threat stimuli for those high in AS, and a study with a subset of this data similarly found AS image LPP uniquely related to self-reported AS (Allan et al., 2019; Saulnier et al., 2021; Taake et al., 2009). A recent review paper even reported similarly increased LPP amplitudes for anxious individuals at intake to unpleasant stimuli (Botelho et al., 2023). Methodological differences aside, there appears to be a replicable positive relationship between AS and LPP. The LPP, an ERP that can be elicited based on individual differences in preference for stimuli (Hajcak et al., 2010), is a physiological marker of motivational salience (Schupp et al., 2000), and it is increased for both negative, or unpleasant, as well as positive, or pleasant, stimuli (Cuthbert et al., 2000). That higher LPP is associated with higher AS in this study suggests that the unpleasant imagery elicited stronger emotional reactivity for those with higher AS, and that the higher amplitude was predictive of greater AS treatment-related change. Together, these findings indicate that AS is associated with increased reactivity unpleasant imagery, and that increase is predictive of treatment outcomes.

The LPP has potential to be explored as a cognitive-affective mechanism underlying Anxiety Sensitivity, as it is sensitive to incoming status for individuals in terms of self-report AS levels. Identifying connections between neurophysiological processes and behavioral expression of AS has immense utility in terms of identifying this reactivity as a treatment target. Identifying empirically-validated referents in the brain to genuine treatment targets allows a more precise and parsimonious understanding of underlying systems. AS is related to a number of different mental health disorders, and our findings of a relationship between AS and the LPP provides a stronger empirical basis for intervention target development. This ERP measure has the ability to offer unique information regarding a sensitivity to AS.

### LPP Related to AS Treatment Change

While several studies have used the P300 and LPP ERPs as a measure of assessing anxiety and mood disorders (Al-Ezzi et al., 2020; Botelho et al., 2023; Hanatani et al., 2005; Miltner et al., 2005; Sahoo, 2016; Zygouris et al., 2022), and our above findings replicate an understanding of an association between ERPs and anxiety and mood disorders, the literature on neural predictors of anxiety treatment response is predominantly comprised of functional magnetic resonance imaging (fMRI) studies (Ball et al., 2014), thus the current research represents novel finding that addresses a critical gap in the literature regarding neural indices of treatment response with EEG/ERP methodology. Importantly though, findings in the current study align with findings from the relative few number of studies that did examine ERPs as predictors of anxiety treatment response, such that higher pretreatment severity was associated with greater treatment response and overall anxiety and mood symptom reduction(Bunford et al., 2017; Kinney et al., 2021; Stange et al., 2017).

Though the studies featured different methodology (using medication as apart of treatment, using cognitive behavioral therapy, or CBT, and/or pediatric sampling), their findings in conjunction with the current study’s findings underlie the potential use for ERPs as neural predictors of treatment response. Anxiety is a globally prevalent mental health problem, and response rates remain inconsistent and low to anxiety and mood disorder treatment(Nimatoudis et al., 2004; Pollack, 2001b). Because AS is associated with the etiology and maintenance of a number of mental health disorders, such as panic disorder, depression, anxiety, and more (Allan et al., 2019), identifying genuine treatment targets is important relative to reaching these afflicted populations on a broader scale. Results from this study replicate findings from emerging research that relates the LPP to anxiety treatment outcome. Validating underlying neural mechanisms is innovative in that it provides a possibility of neuroscience-informed intervention for anxiety and mood pathology. These findings, demonstrating that baseline ERP measures are related to treatment outcome, provide us a deeper understanding of the cognitive-affective neural processes associated with AS.

In line with recent new research on ERPs and treatment response, in the current study, we observed a relationship between the LPP and treatment-related change in AS. Results revealed that, across treatment groups, treatment-related reductions in AS from intake to one month posttreatment were predicted by higher baseline LPP reactivity to unpleasant images, suggesting that those with higher AS at baseline are increasingly responsive to, and benefit more from targeted treatment (Schmidt et al., 2007, 2014, 2017). Further, we found that treatment effects across group were significant relative to the control group. This suggests that LPP measures at intake index reactivity to unpleasant images, which is predictive of treatment-related change, and may represent a target to be further developed. The results from this study are compelling in that LPP activity can not only assist in identifying individuals struggling with elevated AS, but this activity may also be used to then identify people that would benefit from treatment. The interventions in the current study broadly target maladaptive cognitions, and attempt to teach strategies of emotion regulation, thus their higher reactivity relates to better treatment outcome in the sense that they are better able to successfully engage in reappraisal. At baseline, individuals high in AS misinterpret anxiety-related sensations, or higher emotional reactivity, as dangerous, but after treatment, it may be the case that this higher reactivity allows for a more controlled reception and application of regulatory processes to reappraise and lower anxiety symptoms.

The third mind finding is that, related to the second main finding, further analyses revealed evidence of a pattern of separable mechanisms in the CAST and CBM-I groups: the CAST group showed a prediction of treatment outcome parietally, whereas the CBM-I group showed this relationship frontally. The Combined group showed a combined effect in that this condition evidenced treatment outcome predictions in both frontal and parietal areas. The effects are explained by different mechanisms and processes being engaged in the CAST and CBM-I groups, respectively. The CAST group received an intervention focused on dispelling exaggerated anxious thoughts, attending to anxious sensations and fears, and reducing overall emotionality. A main feature of the CAST intervention is exposure, which is known to reduce affective reactivity (Benito et al., 2024; Boswell et al., 2013; Foa & Kozak, 1986). The CAST group condition experienced treatment more focused on emotional reactivity and less on cognitive processing. The main place the increased LPP to emotional stimuli becomes active is parietally (Hajcak et al., 2010), thus individuals in the CAST group that had higher parietal LPP showed greater treatment outcomes because their intervention condition was designed to directly address their reactivity to emotional stimuli. The exposure components targeted a reduction in arousal and overall AS through the CAST group. In contrast, individuals in the CBM-I group received an intervention focused on teaching about negative beliefs, requiring more cognitive processing indicative of frontal activity (Antonenko et al., 2010). CBM-I is designed to directly change cognitive processes like attention and interpretation, thus involves solving tasks that directly target a cognitive bias(MacLeod & Matthews, 2012). Where CAST is meant to address emotionality and reactivity to arousing stimuli, CBM-I is more implicitly cognitive and thought-based, focused not on reducing reactivity to stimuli, but more on actively reinterpreting stimuli in a completely different manner than before.

Altogether, the findings suggest that the P3/LPP is predictive in assessing AS treatment outcomes, and that treatment type plays a role in the cognitive-affective processes that are engaged. CAST interventions that focus on exposure and reduction of anxious sensations and feelings engage the parietal LPP more commonly associated with emotional stimuli reactivity, whereas the CBM-I intervention engages more active thought indicative of frontal activity.

This study identifies the LPP to unpleasant imagery as a vital tool for understanding mechanisms and affective processes involved in AS, an already researched candidate treatment target for anxiety and mood pathology. Increased reactivity to unpleasant images was related to pretreatment AS and posttreatment changes in AS. Given our results, activity within the LPP can assist in identifying individuals struggling with elevated AS, as well as identify individuals that would benefit from treatment. Findings of a relationship between LPP and AS support the notion that AS can be measured physiologically. AS is one of the most well-studied transdiagnostic constructs, and it is associated with the etiology and maintenance of a number of mental health disorders, such as panic disorder, depression, anxiety, and more (Allan et al., 2019). Thus, our findings of a predictive relationship between AS and the LPP provides a stronger empirical basis for intervention target development. P3/LPP is related to salience, motivated attention, and motivational significance. Thus, that this ERP measure can index the amount of unpleasant reactivity offers unique information regarding a sensitivity to AS, thus merging an understanding of psychopathology and treatment with ERP measures could lead to a characterization of separable transdiagnostic systems underlying problem behavior.

Identifying predictors of treatment response is critical, as both patients and clinicians would benefit from objective predictors that indicate which individuals will and will not respond to a specific treatment (Phillips et al., 2000), especially with treatment response rates in anxiety still low (Loerinc et al., 2015; Nimatoudis et al., 2004; Pollack, 2001b; Roth-Rawald et al., 2023). Predictive factors have successfully been identified to predict both mental health service use as well as treatment drop-out rates(DiNapoli et al., 2016; Reneses et al., 2009; Simo et al., 2018), but less research exists relating underlying neural mechanisms as indices of anxiety pathology. As personalized medicine, an approach that favors treatment being viewed as individualized as the disease, rapidly advances in the field of health care (Chan & Ginsburg, 2011), the ability to predict treatment outcome before or shortly after a treatment is initiated could mean significant improvement in individualized treatment selection, and ultimately improvement in the safety and efficacy of treatments. Personalized medicine relies on updating and identifying information to advance understanding of an individual’s vulnerability to certain diseases and disorders (Mathur & Sutton, 2017), and the findings in the current study provide potential to be proactive in tailoring treatments to an individual based on their ERP activity. To uniquely tailor treatment in order for response rates and efficacy to be higher could be transformative in the treating mental health problems afflicting millions broadly.

The patterns we identified regarding within-group correlations of P3/LPP amplitude and AS change scores can inform future research. These findings have high utility in comparing treatment efficacy in order to reduce AS and other INT disorders. Anxiety disorders and symptomatology is widespread issue for millions worldwide (Gustavsson et al., 2011; National Alliance on Mental Illness, 2024; H. U. Wittchen et al., 2011), and there are clearly too few effective treatments available for those afflicted with these INT problems. The potential for an intervention-specific sensitivity to treatment outcome could have important implications for emotion regulation, intervention targets, and interventions broadly. This study’s findings provide an objective neurophysiological marker for AS and opens new opportunities to implement interventions tailored to a specific individual’s needs objectively.

### Limitations

In terms of limitations, although our sample was sufficiently large to detect a relationship between the P3/LPP to unpleasant stimuli and AS treatment response, but power for detecting between-treatment group effects was reduced. Thus, the correlation results showing topographical differences within each respective treatment group, suggesting a unique relationship among treatment type and treatment outcome predicted by ERP amplitude, must be understood more tentatively. Replication, extension with larger samples would be critical to bolster confidence in the reported between-group results.

## Conclusion

In summary, the current study found support for the idea that there exists a sensitivity to AS change scores and different observed treatment types engaged different neural mechanisms. The LPP predicted AS at intake as well as treatment change in AS, and the type of treatment received affects the cognitive-affective processes engaged in treatment outcome. The study further replicates the efficacy of the CAST intervention and illuminates the need for future research on cognitive affect processes as an index for this well-studied transdiagnostic factor of anxiety and mood pathology. Building upon the current work, future research may strive to better integrate this information about the relationship between AS and the physiological domain of ERP measures, with the hope of influencing intervention development.

## Data Availability

All data produced in the present study are available upon reasonable request to the authors.

